# Determinants of early child development among children aged 24–59 months in Ethiopia: 2024–25 Demographic and Health Survey

**DOI:** 10.64898/2026.11.10.26367725

**Authors:** Awoke Mihretu, Mahder Dereb

**Author notes:** Corresponding author: Department of Psychiatry, College of Health Sciences Zambia Street, Tikur Anbessa Specialised Hospital Campus Office No. 601 (CHS Building), P.O. Box 9086, Addis Ababa University Addis Ababa, Ethiopia.

## Abstract

**Background:** The 2024–25 Ethiopia Demographic and Health Survey (EDHS) is the first national survey in Ethiopia to measure early childhood development using UNICEF’s Early Childhood Development Index 2030 (ECDI2030). This provides a national estimate for Sustainable Development Goal (SDG) indicator 4.2.1, the proportion of children aged 24–59 months who are developmentally on track. We estimated the prevalence of being developmentally on track and examined its child, maternal, household and community determinants in Ethiopia.

**Methods:** This cross-sectional study analysed 3,731 children aged 24–59 months (weighted n = 3,514) of 3,232 mothers in 771 clusters. Children were on track if they achieved the age-specific minimum number of the 20 ECDI2030 milestones. Prevalence was estimated with design-based methods. Four three-level mixed-effects logistic regression models (children within mothers within clusters) were fitted, and interviewer effects were assessed.

**Results:** Overall, 36.3% (95% CI 33.4–39.3) of children were developmentally on track. Prevalence ranged from 23.3% in Southwest Regional State, Ethiopia, to 77.7% in Addis Ababa. In the null model, 21.5% of the variation lay between clusters (median odds ratio 3.24). In the final model, the odds of being on track were lower at 36–47 months (adjusted odds ratio [AOR] 0.57, 95% CI 0.44–0.74) and 48–59 months (AOR 0.39, 0.30–0.51) than at 24–35 months. They were also lower after a preceding birth interval under 24 months (AOR 0.75, 0.57–0.99), after recent diarrhoea, fever or cough (AOR 0.59, 0.45–0.77), and where distance to a health facility was a big problem (AOR 0.61, 0.48–0.79). Odds were higher for girls (AOR 1.38, 1.12–1.70), with maternal education (more than secondary vs none: AOR 2.08, 1.19–3.63) and with weekly media exposure (AOR 2.15, 1.51–3.06). The covariates explained 55.9% of the between-cluster variance.

**Conclusions:** Nearly two in three Ethiopian children aged 24–59 months are not developmentally on track, and children fall further behind as they age. Interventions should target maternal education, access to information and health services, birth spacing and childhood illness, with priority for the lowest-performing regions and communities.

## Background

The first years of life are a period of rapid brain development in which motor, language, cognitive, socio-emotional and self-regulatory capacities are built. Development in these years predicts later schooling, health and earnings, and early deficits tend to widen over the life course [1, 2]. An estimated 250 million children under five years in low- and middle-income countries (LMICs) were at risk of not reaching their developmental potential because of stunting or extreme poverty, and this risk is concentrated in sub-Saharan Africa [3, 4]. The Nurturing Care Framework identifies good health, adequate nutrition, responsive caregiving, opportunities for early learning, and security and safety as the inputs young children need [5, 6].

Sustainable Development Goal target 4.2 commits countries to ensuring that all children have access to quality early childhood development, care and pre-primary education. Indicator 4.2.1 is the proportion of children aged 24–59 months who are developmentally on track in health, learning, and psychosocial well-being [7]. Earlier surveys used the 10-item Early Childhood Development Index for children aged 36–59 months, which had ceiling and floor effects and limited content validity. In 2020, UNICEF released the ECDI2030, a 20-item caregiver-reported measure for children aged 24–59 months with age-specific performance standards [7]. ECDI2030 estimates are not comparable with those from the original index.

Ethiopia has adopted a revised policy framework for early childhood development, care and education, along with a national early childhood development strategy [8]. The Ethiopian government has implemented numerous initiatives to improve early childhood development, including daycares in all government offices, community playgrounds, school meals, and other measures. However, population-level evidence on early development remains limited. Multi-country DHS analyses have linked child development to maternal education, household wealth, women’s empowerment and anaemia [9, 10]. A recent analysis of three East African surveys using the ECDI2030 identified child age, maternal education, wealth and access to early learning as leading predictors [11]. Children within the same community share services, infrastructure and norms, and siblings share a caregiver who reports on them. Multilevel models are therefore needed to separate individual from contextual variation [12, 13].

With the 2024–25 EDHS, Ethiopia has, for the first time, nationally representative ECDI2030 data [8]. This study aimed to estimate the prevalence and multilevel determinants of being developmentally on track among Ethiopian children aged 24–59 months.

## Methods

### Study design, setting and data source

This analysis draws on the 2024–25 EDHS, a nationally representative household survey carried out by the Ethiopian Statistical Service with technical support from ICF; fieldwork ran from August 2024 to April 2025 [8]. Households were sampled in two stages. First, 805 enumeration areas were drawn with probability proportional to size within 27 strata formed by crossing region with urban or rural residence; second, 28 households were picked systematically from each area. Security problems prevented fieldwork in eight clusters in Amhara, leaving 797. Interviews were completed in 21,241 of 21,494 occupied households (98.8%) and with 21,395 of 22,231 eligible women aged 15–49 years (96.2%) [8]. Our reporting follows the STROBE checklist for cross-sectional studies [14] (Additional file 3).

### Study population

The ECDI2030 questions sat within the Woman’s Questionnaire and were asked in a random half of the sampled households. In those households, each mother reported on every biological child aged 24–59 months who lived with her [8]. Each ECDI2030 record in the women’s file carries an index (idxecd) pointing to the child’s entry in the birth history, and all 3,731 records matched a living child of the right age. Our sample therefore consisted of 3,731 children (weighted n = 3,514) born to 3,232 mothers in 771 clusters, the same denominator as in the survey report [8]. None of the covariates had missing values.

### Outcome: developmentally on track

The ECDI2030 consists of 20 questions on milestones and behaviours in three domains. The health domain covers gross motor, fine motor and self-care. The learning domain covers expressive language, early literacy, numeracy, pre-writing and executive functioning. The psychosocial domain covers emotional and social skills and internalising and externalising behaviour [7]. Items 1–18 are yes/no questions and were scored 1 for “yes”; “no” and “don’t know” were scored 0. Item 19, how often the child seems very sad or depressed, was scored 1 unless the answer was “daily” or “don’t know”. Item 20, how much the child kicks, bites or hits other children compared with peers, was scored 1 for “not at all” or “the same or less” [7]. A child was classified as developmentally on track if the summed score reached at least 7 (24–29 months), 9 (30–35), 11 (36–41), 13 (42–47) or 15 (48–59 months) [7, 8]. Because the ECDI2030 is designed to give a single summary indicator, domain-specific estimates were not computed; item-level results are shown in Additional file 1. Our coding reproduced the published national estimate (36.3%) and all published subgroup estimates exactly [8].

### Covariates

Covariate variables were selected a priori from the Nurturing Care Framework and previous DHS-based studies [5, 9–11].

**Child-level** variables were age (24–35, 36–47, 48–59 months), sex, birth order (1, 2–3, 4–5, ≥6), a preceding birth interval of less than 24 months, receipt of vitamin A in the past six months, and diarrhoea, fever or cough in the two weeks before the survey.

**Maternal** variables were age (15–24, 25–34, 35–49 years), highest education, marital status, current work, at least weekly exposure to a newspaper, radio or television, and ever use of the internet.

**Household and community** variables were sex of the household head, household size (≤4, 5–7, ≥8 members), wealth quintile, whether distance to a health facility was a big problem when seeking care, place of residence and region. “Don’t know” responses for vitamin A and illness were coded as “no”. Anthropometric measurements were taken in the other half of households and were therefore unavailable for these children.

### Statistical analysis

All analyses were carried out in Stata version 18 [15]. For national and subgroup prevalence, we declared the survey design (svyset) using the women’s sampling weight, the 771 clusters as primary sampling units and the 27 sampling strata. Standard errors came from Taylor linearisation, and 95% confidence intervals (CI) were computed on the logit scale. Differences in prevalence across categories were judged with the design-adjusted Wald F test.

To study determinants, we treated the data as three nested levels, children (level 1) within mothers (level 2) within clusters (level 3), and fitted random-intercept logistic models with melogit, using mean–variance adaptive Gauss–Hermite quadrature with 25 integration points. The models were built in four steps: an empty model (Model 0); child and maternal characteristics (Model 1); household and community characteristics alone (Model 2); and all characteristics together (Model 3). Region and residence, which define the sampling strata, were entered as fixed effects, and each covariate was also fitted on its own to give crude odds ratios (COR). Clustering was summarised with the intraclass correlation (ICC = σ²(cluster)/[σ²(cluster) + σ²(mother) + π²/3]); the cluster variance was expressed as a median odds ratio (MOR = exp[0.954√σ²(cluster)]); and the share of it explained by each set of covariates was given by the proportional change in variance (PCV) [12, 13]. We retained the model with the lowest Akaike information criterion (AIC), using the Bayesian information criterion and log-likelihood as secondary guides. Because level-specific weights are not released with DHS data, the multilevel models were fitted without weights, as discussed in DHS methodological guidance [16]; as a check, we also fitted a design-weighted single-level logistic regression, which gives population-averaged odds ratios. Associations are reported as adjusted odds ratios (AOR) with 95% CIs, and p < 0.05 was taken as statistically significant.

ECDI2030 answers come from mothers but are recorded by interviewers, so differences in how interviewers asked or coded the questions could look like community variation. We examined this in two ways. First, we compared a two-level model with a cluster random intercept with one that had an interviewer random intercept instead. Second, for each of the 161 interviewers, we compared the number of children recorded as on track with the number expected from colleagues working in the same clusters (Mantel–Haenszel test stratified by cluster), flagging interviewers whose two-sided p-value fell below the Bonferroni threshold (0.05/161). The full model was then refitted without the children these interviewers had assessed.

Claude (Anthropic), an AI model, helped with data management and with writing the analysis code; we checked every step and every result ourselves. The complete Stata code is given in Additional file 2.

#### Ethical considerations

We used de-identified data that the DHS Program makes available to registered researchers, and we had no contact with survey participants. The DHS Program approved our request to use the 2024–25 EDHS datasets, which we downloaded from https://dhsprogram.com/data/available-datasets.cfm. Because this was a secondary analysis, we did not seek further ethical review. The survey itself was approved by the Ethiopian Public Health Institute Institutional Review Board and the ICF Institutional Review Board, and every respondent gave informed consent before being interviewed [8].

## Results

### Characteristics of the study population

Of the 3,731 children (weighted n = 3,514), 52.7% were boys and 72.4% lived in rural areas. Nearly half (45.8%) had mothers with no formal education, 27.9% had mothers with weekly media exposure and 10.0% had mothers who had used the internet. One in five (20.7%) were born less than 24 months after a previous birth, and 19.9% had diarrhoea, fever or cough in the previous two weeks (Table 1).

**Table 1.** Characteristics of children aged 24–59 months and weighted prevalence of being developmentally on track (ECDI2030), Ethiopia DHS 2024–25 (n = 3,731)

| **Characteristic** | **Unweighted n** | **Weighted %** | **% on track (95% CI)** | **p-value<sup>a</sup>** |
|---|---|---|---|---|
| **Child characteristics** |  |  |  |  |
| Child’s age (months) |  |  |  | 0.007 |
| 24–35 | 1,157 | 31.0 | 42.6 (37.8–47.6) |  |
| 36–47 | 1,311 | 36.1 | 33.6 (29.3–38.2) |  |
| 48–59 | 1,263 | 32.9 | 33.4 (29.1–37.9) |  |
| Child’s sex |  |  |  | 0.248 |
| Male | 1,964 | 52.7 | 35.1 (31.7–38.6) |  |
| Female | 1,767 | 47.3 | 37.7 (33.8–41.8) |  |
| Birth order |  |  |  | 0.006 |
| 1 | 808 | 22.6 | 41.8 (36.3–47.6) |  |
| 2–3 | 1,302 | 36.4 | 38.6 (34.5–42.8) |  |
| 4–5 | 841 | 21.7 | 31.1 (26.3–36.4) |  |
| ≥6 | 780 | 19.3 | 31.5 (26.3–37.3) |  |
| Preceding birth interval |  |  |  | 0.012 |
| ≥24 months or first birth | 2,906 | 79.3 | 37.9 (34.6–41.3) |  |
| <24 months | 825 | 20.7 | 30.2 (25.2–35.8) |  |
| Vitamin A in last 6 months |  |  |  | 0.072 |
| No | 2,328 | 64.0 | 34.4 (30.7–38.3) |  |
| Yes | 1,403 | 36.0 | 39.7 (35.3–44.4) |  |
| Diarrhoea, fever or cough in last 2 weeks |  |  |  | 0.979 |
| No | 2,865 | 80.1 | 36.3 (33.1–39.6) |  |
| Yes | 866 | 19.9 | 36.4 (31.3–41.8) |  |
| **Maternal characteristics** |  |  |  |  |
| Mother’s age (years) |  |  |  | 0.264 |
| 15–24 | 620 | 15.5 | 37.8 (30.7–45.5) |  |
| 25–34 | 2,007 | 53.5 | 37.8 (34.1–41.6) |  |
| 35–49 | 1,104 | 31.1 | 33.0 (28.3–38.2) |  |
| Mother’s education |  |  |  | <0.001 |
| No education | 1,773 | 45.8 | 28.6 (24.8–32.8) |  |
| Primary | 1,228 | 34.9 | 38.4 (33.5–43.6) |  |
| Secondary | 428 | 12.0 | 43.2 (35.3–51.5) |  |
| More than secondary | 302 | 7.3 | 62.9 (52.1–72.6) |  |
| Marital status |  |  |  | 0.883 |
| Married/in union | 3,463 | 94.5 | 36.4 (33.4–39.4) |  |
| Not in union | 268 | 5.5 | 35.4 (23.4–49.5) |  |
| Mother currently working |  |  |  | 0.122 |
| No | 2,592 | 63.3 | 34.6 (31.0–38.3) |  |
| Yes | 1,139 | 36.7 | 39.3 (34.6–44.2) |  |
| Weekly exposure to newspaper, radio or TV |  |  |  | <0.001 |
| No | 2,904 | 72.1 | 29.3 (26.2–32.6) |  |
| Yes | 827 | 27.9 | 54.5 (48.8–60.1) |  |
| Mother ever used the internet |  |  |  | <0.001 |
| No | 3,315 | 90.0 | 33.1 (30.2–36.2) |  |
| Yes | 416 | 10.0 | 64.9 (56.1–72.8) |  |
| **Household and community characteristics** |  |  |  |  |
| Sex of household head |  |  |  | 0.014 |
| Male | 2,918 | 84.5 | 34.6 (31.4–38.0) |  |
| Female | 813 | 15.5 | 45.5 (38.0–53.2) |  |
| Household size (members) |  |  |  | 0.188 |
| ≤4 | 1,015 | 28.3 | 40.0 (35.6–44.7) |  |
| 5–7 | 1,773 | 50.8 | 34.5 (30.5–38.8) |  |
| ≥8 | 943 | 20.9 | 35.6 (30.2–41.4) |  |
| Wealth quintile |  |  |  | <0.001 |
| Lowest | 1,413 | 23.4 | 27.2 (22.0–33.0) |  |
| Second | 720 | 21.2 | 29.3 (24.2–35.1) |  |
| Middle | 489 | 17.7 | 34.4 (28.3–41.1) |  |
| Fourth | 492 | 19.8 | 40.4 (33.6–47.5) |  |
| Highest | 617 | 18.0 | 53.9 (46.2–61.3) |  |
| Distance to health facility |  |  |  | <0.001 |
| Not a big problem | 1,972 | 54.4 | 41.2 (37.0–45.6) |  |
| Big problem | 1,759 | 45.6 | 30.5 (26.4–34.8) |  |
| Residence |  |  |  | <0.001 |
| Urban | 1,041 | 27.6 | 50.8 (45.1–56.5) |  |
| Rural | 2,690 | 72.4 | 30.8 (27.5–34.2) |  |
| Region |  |  |  | <0.001 |
| Tigray | 299 | 5.9 | 30.5 (24.8–36.9) |  |
| Afar | 364 | 1.4 | 34.3 (25.4–44.4) |  |
| Amhara | 254 | 21.7 | 29.0 (23.8–34.7) |  |
| Oromia | 370 | 39.6 | 34.8 (28.8–41.2) |  |
| Somali | 467 | 5.2 | 45.8 (38.7–53.0) |  |
| Benishangul-Gumuz | 220 | 1.1 | 30.4 (22.0–40.2) |  |
| Central Ethiopia | 294 | 5.1 | 43.3 (37.2–49.6) |  |
| Sidama | 237 | 3.5 | 46.0 (38.3–53.9) |  |
| South West Ethiopia | 221 | 2.7 | 23.3 (17.4–30.4) |  |
| South Ethiopia | 320 | 8.5 | 36.3 (28.9–44.5) |  |
| Gambella | 210 | 0.5 | 36.3 (29.3–43.9) |  |
| Harari | 170 | 0.3 | 62.3 (53.4–70.5) |  |
| Addis Ababa | 159 | 4.1 | 77.7 (69.9–84.0) |  |
| Dire Dawa | 146 | 0.4 | 49.1 (38.7–59.6) |  |
| Total | 3,731 | 100.0 | 36.3 (33.4–39.3) |  |

Weighted percentages and prevalence account for sampling weights, clustering and stratification.
<sup>a</sup> Design-adjusted Wald F test.

### Prevalence of being developmentally on track

Overall, 36.3% (95% CI 33.4–39.3) of children were developmentally on track (Table 1). Prevalence was 42.6% at 24–35 months and about one third at 36–47 (33.6%) and 48–59 months (33.4%; p = 0.007). Milestones in early literacy and numeracy were achieved by few children, even at 48–59 months: 26.5% recognised five letters, 20.2% recognised the numbers 1–5 and 8.7% could write their name (Additional file 1, Table S1). Prevalence rose with mother’s education, from 28.6% (no education) to 62.9% (more than secondary), and with household wealth, from 27.2% in the lowest to 53.9% in the highest quintile. It was higher among children whose mothers had weekly media exposure (54.5% vs 29.3%) or had used the internet (64.9% vs 33.1%), and in urban than in rural areas (50.8% vs 30.8%). Prevalence was lower where distance to a health facility was a big problem (30.5% vs 41.2%) and after a short preceding birth interval (30.2% vs 37.9%). By region, it ranged from 23.3% in Southwest Regional State and 29.0% in Amhara Regional State to 62.3% in Harari Regional State and 77.7% in Addis Ababa City Administration.

### Between-cluster variation and model fit

In the null model, the variance was 1.52 between clusters and 2.25 between mothers. This gave a cluster ICC of 21.5%, a sibling correlation of 53.4% and a MOR of 3.24 (Table 2). A two-level model fitted much better than a single-level model (likelihood-ratio χ² = 152, p < 0.001). Child and maternal variables reduced the cluster variance by 32.9% (Model 1) and household and community variables by 54.0% (Model 2). The full model (Model 3) explained 55.9% of the cluster variance, had the lowest AIC (4,472) and was retained. Residual clustering remained substantial (ICC 10.8%; MOR 2.18).

**Table 2.**
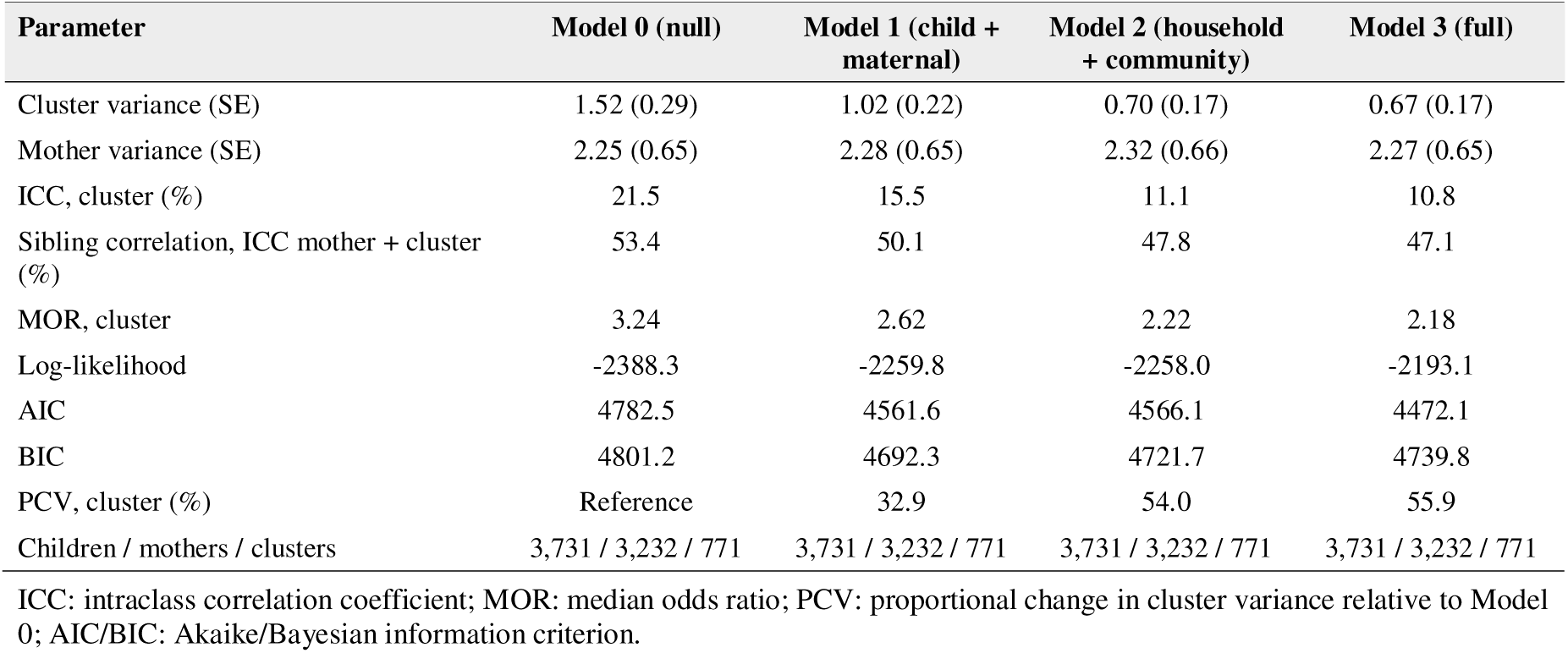
Random-effects parameters and model fit of the three-level logistic regression models for being developmentally on track, Ethiopia DHS 2024–25

### Factors associated with being developmentally on track

In the final model (Table 3), the odds of being on track were lower among children aged 36–47 months (AOR 0.57, 95% CI 0.44–0.74) and 48–59 months (AOR 0.39, 0.30–0.51) than among those aged 24–35 months. Girls had higher odds than boys (AOR 1.38, 1.12–1.70). Odds were lower among children born less than 24 months after a previous birth (AOR 0.75, 0.57–0.99) and those with diarrhoea, fever or cough in the previous two weeks (AOR 0.59, 0.45–0.77). Compared with children of mothers with no education, odds were higher for those whose mothers had primary (AOR 1.37, 1.02–1.82), secondary (AOR 1.86, 1.22–2.83) or more than secondary education (AOR 2.08, 1.19–3.63). Maternal weekly media exposure was associated with higher odds (AOR 2.15, 1.51–3.06). Children in households where distance to a health facility was a big problem had lower odds (AOR 0.61, 0.48–0.79), and those in the middle wealth quintile had higher odds than the poorest (AOR 1.62, 1.09–2.41). Compared with Addis Ababa, odds were lower in most regions, most markedly in South West Ethiopia (AOR 0.12, 0.05–0.29), Amhara (AOR 0.16, 0.07–0.36), Benishangul-Gumuz (AOR 0.17, 0.07–0.41) and Tigray (AOR 0.18, 0.08–0.40); Harari, Somali and Sidama did not differ significantly. Large crude associations with residence (COR 0.26), wealth (highest vs lowest COR 8.13) and internet use (COR 5.62) were largely explained by the other variables. Birth order, vitamin A, mother’s age, marital status and work, sex of household head and household size were not independently associated.

**Table 3.**
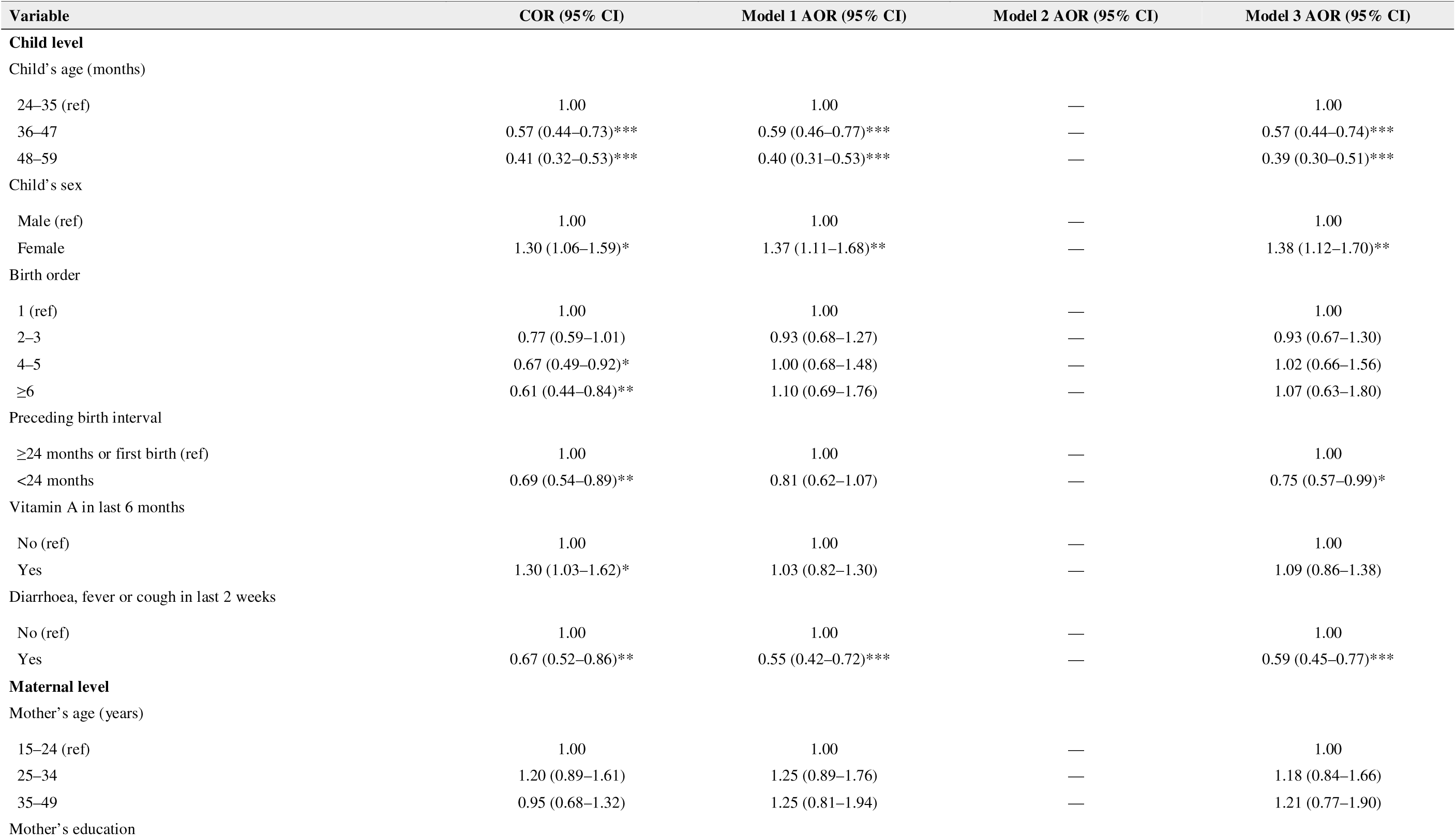
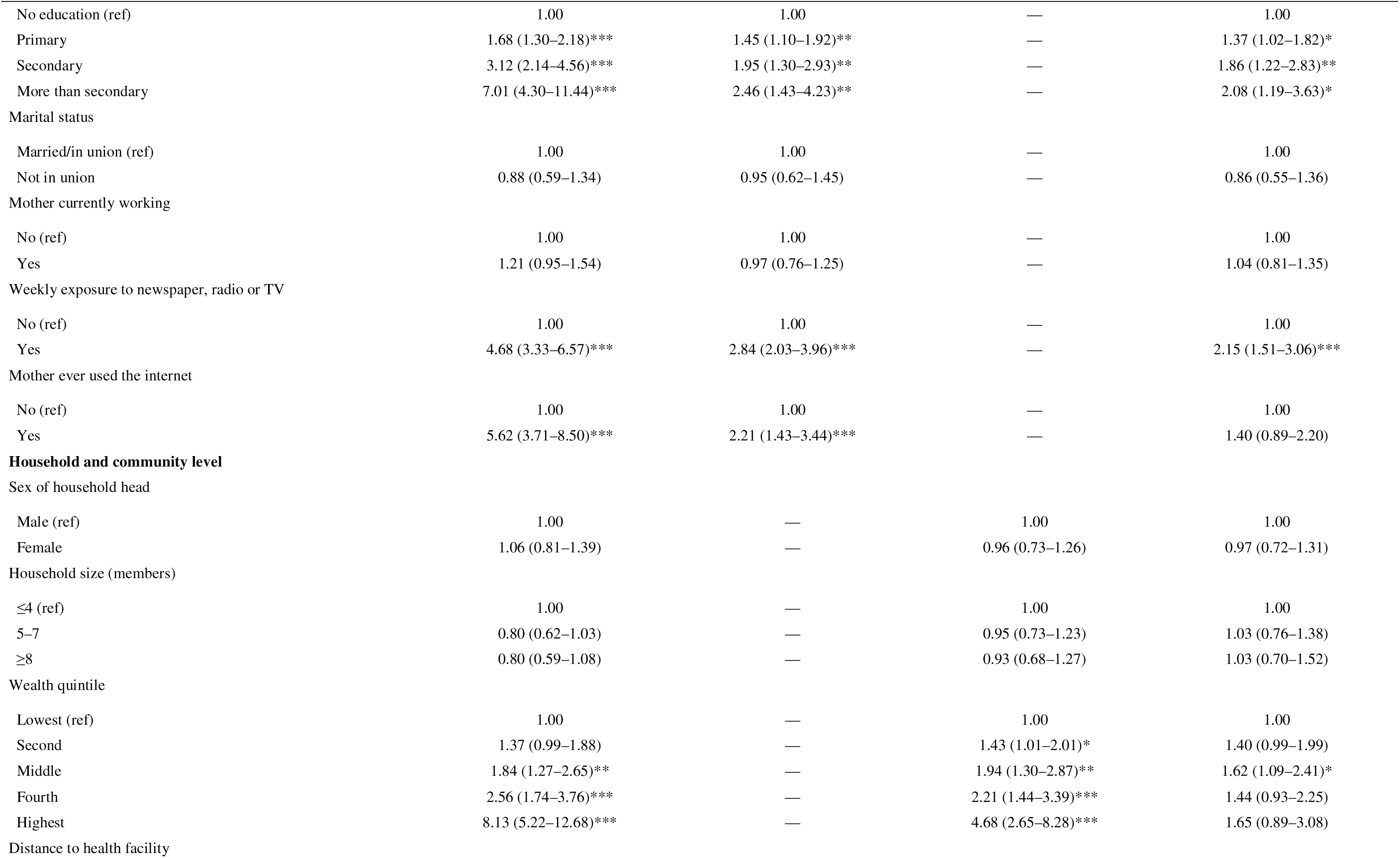
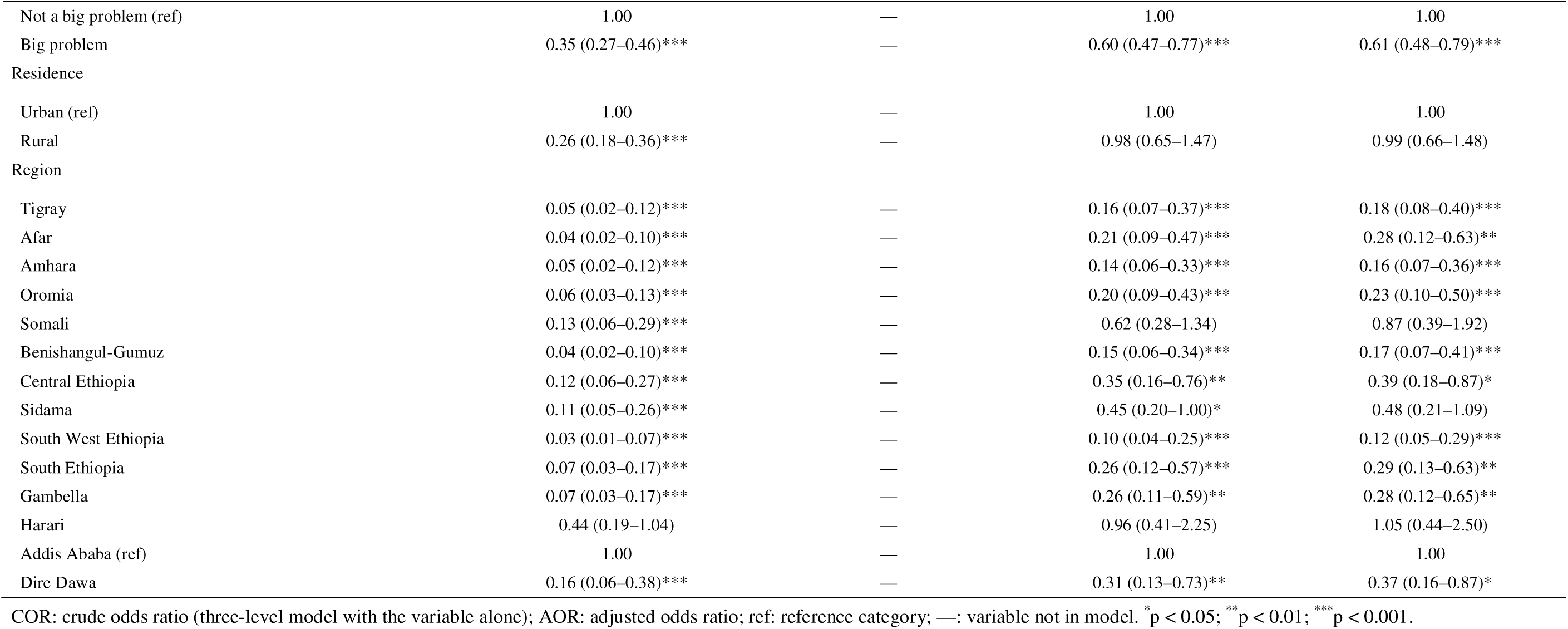
Factors associated with being developmentally on track among children aged 24–59 months: three-level mixed-effects logistic regression (children hin mothers within clusters), Ethiopia DHS 2024–25 (n = 3,731)

| Variable | COR (95% CI) | Model 1 AOR (95% CI) | Model 2 AOR (95% CI) | Model 3 AOR (95% CI) |
|---|---|---|---|---|
| **Child level** |  |  |  |  |
| Child’s age (months) |  |  |  |  |
| 24–35 (ref) | 1.00 | 1.00 | — | 1.00 |
| 36–47 | 0.57 (0.44–0.73)*** | 0.59 (0.46–0.77)*** | — | 0.57 (0.44–0.74)*** |
| 48–59 | 0.41 (0.32–0.53)*** | 0.40 (0.31–0.53)*** | — | 0.39 (0.30–0.51)*** |
| Child’s sex |  |  |  |  |
| Male (ref) | 1.00 | 1.00 | — | 1.00 |
| Female | 1.30 (1.06–1.59)* | 1.37 (1.11–1.68)** | — | 1.38 (1.12–1.70)** |
| Birth order |  |  |  |  |
| 1 (ref) | 1.00 | 1.00 | — | 1.00 |
| 2–3 | 0.77 (0.59–1.01) | 0.93 (0.68–1.27) | — | 0.93 (0.67–1.30) |
| 4–5 | 0.67 (0.49–0.92)* | 1.00 (0.68–1.48) | — | 1.02 (0.66–1.56) |
| ≥6 | 0.61 (0.44–0.84)** | 1.10 (0.69–1.76) | — | 1.07 (0.63–1.80) |
| Preceding birth interval |  |  |  |  |
| ≥24 months or first birth (ref) | 1.00 | 1.00 | — | 1.00 |
| <24 months | 0.69 (0.54–0.89)** | 0.81 (0.62–1.07) | — | 0.75 (0.57–0.99)* |
| Vitamin A in last 6 months |  |  |  |  |
| No (ref) | 1.00 | 1.00 | — | 1.00 |
| Yes | 1.30 (1.03–1.62)* | 1.03 (0.82–1.30) | — | 1.09 (0.86–1.38) |
| Diarrhoea, fever or cough in last 2 weeks |  |  |  |  |
| No (ref) | 1.00 | 1.00 | — | 1.00 |
| Yes | 0.67 (0.52–0.86)** | 0.55 (0.42–0.72)*** | — | 0.59 (0.45–0.77)*** |
| **Maternal level** |  |  |  |  |
| Mother’s age (years) |  |  |  |  |
| 15–24 (ref) | 1.00 | 1.00 | — | 1.00 |
| 25–34 | 1.20 (0.89–1.61) | 1.25 (0.89–1.76) | — | 1.18 (0.84–1.66) |
| 35–49 | 0.95 (0.68–1.32) | 1.25 (0.81–1.94) | — | 1.21 (0.77–1.90) |
| Mother’s education |  |  |  |  |
| No education (ref) | 1.00 | 1.00 | — | 1.00 |
| Primary | 1.68 (1.30–2.18)*** | 1.45 (1.10–1.92)** | — | 1.37 (1.02–1.82)* |
| Secondary | 3.12 (2.14–4.56)*** | 1.95 (1.30–2.93)** | — | 1.86 (1.22–2.83)** |
| More than secondary | 7.01 (4.30–11.44)*** | 2.46 (1.43–4.23)** | — | 2.08 (1.19–3.63)* |
| Marital status |  |  |  |  |
| Married/in union (ref) | 1.00 | 1.00 | — | 1.00 |
| Not in union | 0.88 (0.59–1.34) | 0.95 (0.62–1.45) | — | 0.86 (0.55–1.36) |
| Mother currently working |  |  |  |  |
| No (ref) | 1.00 | 1.00 | — | 1.00 |
| Yes | 1.21 (0.95–1.54) | 0.97 (0.76–1.25) | — | 1.04 (0.81–1.35) |
| Weekly exposure to newspaper, radio or TV |  |  |  |  |
| No (ref) | 1.00 | 1.00 | — | 1.00 |
| Yes | 4.68 (3.33–6.57)*** | 2.84 (2.03–3.96)*** | — | 2.15 (1.51–3.06)*** |
| Mother ever used the internet |  |  |  |  |
| No (ref) | 1.00 | 1.00 | — | 1.00 |
| Yes | 5.62 (3.71–8.50)*** | 2.21 (1.43–3.44)*** | — | 1.40 (0.89–2.20) |
| **Household and community level** |  |  |  |  |
| Sex of household head |  |  |  |  |
| Male (ref) | 1.00 | — | 1.00 | 1.00 |
| Female | 1.06 (0.81–1.39) | — | 0.96 (0.73–1.26) | 0.97 (0.72–1.31) |
| Household size (members) |  |  |  |  |
| ≤4 (ref) | 1.00 | — | 1.00 | 1.00 |
| 5–7 | 0.80 (0.62–1.03) | — | 0.95 (0.73–1.23) | 1.03 (0.76–1.38) |
| ≥8 | 0.80 (0.59–1.08) | — | 0.93 (0.68–1.27) | 1.03 (0.70–1.52) |
| Wealth quintile |  |  |  |  |
| Lowest (ref) | 1.00 | — | 1.00 | 1.00 |
| Second | 1.37 (0.99–1.88) | — | 1.43 (1.01–2.01)* | 1.40 (0.99–1.99) |
| Middle | 1.84 (1.27–2.65)** | — | 1.94 (1.30–2.87)** | 1.62 (1.09–2.41)* |
| Fourth | 2.56 (1.74–3.76)*** | — | 2.21 (1.44–3.39)*** | 1.44 (0.93–2.25) |
| Highest | 8.13 (5.22–12.68)*** | — | 4.68 (2.65–8.28)*** | 1.65 (0.89–3.08) |
| Distance to health facility |  |  |  |  |
| Not a big problem (ref) | 1.00 | — | 1.00 | 1.00 |
| Big problem | 0.35 (0.27–0.46)*** | — | 0.60 (0.47–0.77)*** | 0.61 (0.48–0.79)*** |
| Residence |  |  |  |  |
| Urban (ref) | 1.00 | — | 1.00 | 1.00 |
| Rural | 0.26 (0.18–0.36)*** | — | 0.98 (0.65–1.47) | 0.99 (0.66–1.48) |
| Region |  |  |  |  |
| Tigray | 0.05 (0.02–0.12)*** | — | 0.16 (0.07–0.37)*** | 0.18 (0.08–0.40)*** |
| Afar | 0.04 (0.02–0.10)*** | — | 0.21 (0.09–0.47)*** | 0.28 (0.12–0.63)** |
| Amhara | 0.05 (0.02–0.12)*** | — | 0.14 (0.06–0.33)*** | 0.16 (0.07–0.36)*** |
| Oromia | 0.06 (0.03–0.13)*** | — | 0.20 (0.09–0.43)*** | 0.23 (0.10–0.50)*** |
| Somali | 0.13 (0.06–0.29)*** | — | 0.62 (0.28–1.34) | 0.87 (0.39–1.92) |
| Benishangul-Gumuz | 0.04 (0.02–0.10)*** | — | 0.15 (0.06–0.34)*** | 0.17 (0.07–0.41)*** |
| Central Ethiopia | 0.12 (0.06–0.27)*** | — | 0.35 (0.16–0.76)** | 0.39 (0.18–0.87)* |
| Sidama | 0.11 (0.05–0.26)*** | — | 0.45 (0.20–1.00)* | 0.48 (0.21–1.09) |
| South West Ethiopia | 0.03 (0.01–0.07)*** | — | 0.10 (0.04–0.25)*** | 0.12 (0.05–0.29)*** |
| South Ethiopia | 0.07 (0.03–0.17)*** | — | 0.26 (0.12–0.57)*** | 0.29 (0.13–0.63)** |
| Gambella | 0.07 (0.03–0.17)*** | — | 0.26 (0.11–0.59)** | 0.28 (0.12–0.65)** |
| Harari | 0.44 (0.19–1.04) | — | 0.96 (0.41–2.25) | 1.05 (0.44–2.50) |
| Addis Ababa (ref) | 1.00 | — | 1.00 | 1.00 |
| Dire Dawa | 0.16 (0.06–0.38)*** | — | 0.31 (0.13–0.73)** | 0.37 (0.16–0.87)* |

COR: crude odds ratio (three-level model with the variable alone); AOR: adjusted odds ratio; ref: reference category; —: variable not in model. <sup>\*</sup>p < 0.05; <sup>\*\*</sup>p < 0.01; <sup>\*\*\*</sup>p < 0.001.

### Interviewer effects and sensitivity analyses

Replacing the cluster random intercept with an interviewer random intercept improved model fit (AIC 4,197 vs 4,502) and raised the ICC from 12.1% to 25.2%, so some of the apparent clustering reflects differences between interviewers rather than between communities. Seven of 161 interviewers were flagged: four recorded more and three fewer children on track than other interviewers in the same clusters (Additional file 1, Table S3). These seven interviewers had interviewed 2.7% of the weighted sample. Excluding them changed the national prevalence only slightly, to 35.8% (32.8–38.9). Associations with age, sex, recent illness, maternal education and media exposure were unchanged; those with distance to a health facility (AOR 0.80, 0.62–1.02) and short birth interval (AOR 0.79, 0.60–1.04) were weaker and no longer significant, whereas internet use (AOR 1.60, 1.01–2.52) and the highest wealth quintile (AOR 1.88, 1.01–3.51) became significant (Additional file 1, Table S2). In design-based single-level models, the associations with age and media exposure remained significant, whereas those with maternal education, recent illness and distance were smaller and not significant, as expected for population-averaged estimates.

## Discussion

This study provides the first national estimate of SDG indicator 4.2.1 for Ethiopia using the ECDI2030, and the first multilevel analysis of its determinants. Only about one in three children aged 24–59 months (36.3%) was developmentally on track. Children were more likely to be on track if they were younger, female, born after an adequate birth interval and recently free of illness, if their mothers were educated and exposed to media, if their households were within reach of health services, and if they lived in better-performing regions. Development was strongly clustered within families and communities, and partly within interviewers.

The national prevalence is similar to recent ECDI2030 estimates for Mozambique (39%) and lower than those for Tanzania (47%) and Kenya (78%) [11]. Because the ECDI2030 uses different items, ages and scoring from the original index, these estimates should not be compared with earlier figures based on the 10-item ECDI or used to infer trends [7].

The age-specific standards are designed so that a similar share of children would be on track at each age in a reference population. The decline from 42.6% at 24–35 months to about one third at older ages, and the strong age gradient after adjustment, therefore suggest that Ethiopian children fall further behind as expectations shift towards language, early literacy and numeracy. Item-level data support this: at 48–59 months, only about a quarter of children recognised letters or counted to ten, and fewer than one in ten could write their name. These findings point to the importance of early learning opportunities at home and of expanding pre-primary education before school entry [1, 2, 8].

Maternal education showed a clear gradient that persisted after adjustment for wealth, residence and region, in agreement with multi-country evidence [3, 9, 17]. Educated mothers are more likely to provide responsive care and learning opportunities and to use health services [2, 17]. Weekly exposure to media was among the strongest independent correlates. It may provide parenting and health information, but in cross-sectional data it also marks household resources and engagement, so it should not be interpreted as causal. The large crude wealth and urban–rural gradients were mostly explained by maternal education, media exposure, access to services and region. This suggests that these more proximal factors carry much of the socioeconomic disadvantage.

Recent illness and short birth intervals were independently associated with lower odds of being on track. Both are consistent with the Nurturing Care Framework, in which good health and adequate caregiver time and resources support development [5]. Recent illness may reduce a child’s activity and opportunities for learning, and may also mark recurrent illness and undernutrition, which we could not measure. The association with illness was seen only in comparisons within communities and families: weighted prevalence did not differ by recent illness (36.4% vs 36.3%), and the population-averaged odds ratio (0.80) was not significant, so this finding should be interpreted with caution. Short birth intervals may dilute maternal attention and household resources. Distance to health facilities, a marker of service access, was also associated with poorer development, although this association weakened when outlying interviewers were excluded, which supports integrating developmental monitoring and caregiver counselling into routine child health contacts and the Health Extension Programme. Girls had higher odds of being on track than boys, consistent with sex differences in early language and social development reported in other settings.

Regional differences remained large after adjustment. South West Ethiopia, Amhara, Benishangul-Gumuz and Tigray had the lowest odds; Amhara and Tigray have been affected by conflict, and South West Ethiopia and Benishangul-Gumuz have limited services. Somali and Sidama had higher prevalence than expected from their socioeconomic profile. This may reflect real differences in caregiving but could also reflect how milestones were interpreted and reported in different languages; measurement invariance of the ECDI2030 across Ethiopian languages has not been established. The substantial interviewer variance supports this caution. Interviewer effects did not bias the national estimate, which changed by less than one percentage point when outlying interviewers were excluded, but they add uncertainty to comparisons between small areas. Residual between-cluster variation (MOR 2.18) indicates that unmeasured community factors, such as availability of pre-primary classes, quality of health services, conflict exposure and child-rearing norms, also matter. This supports area-based targeting of nurturing-care interventions.

### Strengths and limitations

This study has several strengths. It uses the most recent nationally representative data, applies the internationally agreed SDG 4.2.1 measure, reproduces the official estimates exactly, separates family from community variation with three-level models, and checks explicitly for interviewer effects. It also has limitations. Being cross-sectional, it cannot establish causation, and covariates such as recent illness and media exposure may stand in for other, unmeasured factors. The ECDI2030 relies on what mothers report and is designed to describe populations, not to identify individual children with developmental delay [7]. Children who were not living with their mothers were not assessed. We could not adjust for nutritional status or home stimulation, which were not measured for these children, or for early childhood education, which was recorded only from age 4 years. Finally, the multilevel models were unweighted, and some associations were weaker in the population-averaged models.

## Conclusions

Only about one in three Ethiopian children aged 24–59 months is developmentally on track, and children fall further behind as they approach school age. Maternal education, media exposure, access to health services, adequate birth spacing and freedom from illness are associated with better development, and large regional and community differences remain. Achieving SDG target 4.2 will require scaling up nurturing-care and early-learning interventions, with priority for less-educated households, areas far from health services and the lowest-performing regions.

## Data Availability

All the data is publicly available at https://dhsprogram.com/data.

## Abbreviations

AIC: Akaike information criterion
AOR: adjusted odds ratio
CI: confidence interval
COR: crude odds ratio
DHS: Demographic and Health Survey
ECDI2030: Early Childhood Development Index 2030
EDHS: Ethiopia Demographic and Health Survey
ICC: intraclass correlation coefficient
MOR: median odds ratio
PCV: proportional change in variance
SDG: Sustainable Development Goal.

## Declarations

### Ethics approval and consent to participate

Not applicable. We analysed de-identified secondary data with the permission of the DHS Program and had no contact with participants.

The 2024–25 EDHS was approved by the Ethiopian Public Health Institute Institutional Review Board and the ICF Institutional Review Board, and the survey team obtained informed consent from all respondents [8].

## Consent for publication

Not applicable.

## Responsible AI use

Claude (Anthropic) was used to support data management, coding and the preparation of draft text. The authors checked all outputs against the source data and the survey report and are responsible for the content of this article.

## Availability of data and materials

The 2024–25 EDHS datasets can be requested free of charge from the DHS Program (https://dhsprogram.com). The Stata code used for this analysis is provided in Additional file 2.

## Authors’ contributions

AM conceived the study and drafted the manuscript. MD conducted the data analysis and interpretation. Both authors read and approved the final manuscript.

## Acknowledgements

We are grateful to the Ethiopian Statistical Service and the DHS Program for making the data available, and to the mothers who shared information about their children.

---

**Additional file 1, Table S1.** Weighted percentage of children achieving each ECDI2030 milestone, by age group, Ethiopia DHS 2024–25

| **Item** | **Domain** | **24–35 months** | **36–47 months** | **48–59 months** | **All** |
|---|---|---|---|---|---|
| 1. Walks on an uneven surface without falling | Health | 55.9 | 65.3 | 80.0 | 67.2 |
| 2. Jumps with both feet | Health | 52.8 | 70.3 | 83.7 | 69.3 |
| 3. Dresses self without help | Health | 22.6 | 46.8 | 71.8 | 47.6 |
| 4. Fastens and unfastens buttons | Health | 16.9 | 38.5 | 66.7 | 41.1 |
| 5. Says 10 or more words | Learning | 64.6 | 70.8 | 83.0 | 72.9 |
| 6. Speaks in 3-word sentences | Learning | 57.0 | 74.7 | 84.0 | 72.3 |
| 7. Speaks in 5-word sentences | Learning | 31.1 | 57.9 | 75.2 | 55.3 |
| 8. Uses I/you/she/he correctly | Learning | 46.2 | 68.4 | 82.4 | 66.1 |
| 9. Names a familiar object | Learning | 46.6 | 66.6 | 79.3 | 64.6 |
| 10. Recognises 5+ letters | Learning | 5.7 | 13.5 | 26.5 | 15.4 |
| 11. Writes own name | Learning | 0.9 | 1.6 | 8.7 | 3.7 |
| 12. Recognises numbers 1–5 | Learning | 2.4 | 7.0 | 20.2 | 9.9 |
| 13. Gives 3 objects when asked | Learning | 7.8 | 22.5 | 43.8 | 24.9 |
| 14. Counts 10 objects | Learning | 3.8 | 11.5 | 24.6 | 13.4 |
| 15. Persists in activity without help | Learning | 8.3 | 17.7 | 28.5 | 18.4 |
| 16. Asks about familiar people | Psychosocial | 24.8 | 41.5 | 58.3 | 41.9 |
| 17. Offers to help | Psychosocial | 30.2 | 42.6 | 57.8 | 43.7 |
| 18. Gets along with other children | Psychosocial | 60.2 | 70.7 | 80.4 | 70.6 |
| 19. Not sad/depressed daily | Psychosocial | 79.8 | 80.7 | 86.0 | 82.2 |
| 20. Kicks/bites/hits no more than peers | Psychosocial | 72.1 | 71.5 | 75.0 | 72.8 |
| Mean number of milestones achieved |  | 6.9 | 9.4 | 12.2 |  |
| Age-specific threshold for “on track” |  | 7 / 9 (24–29 / 30–35) | 11 / 13 (36–41 / 42–47) | 15 |  |

Items 1–18: “yes”; item 19: sad or depressed less often than daily; item 20: kicks, bites or hits not at all or the same or less than peers. “Don’t know” scored as not achieved.

**Additional file 1, Table S2.**
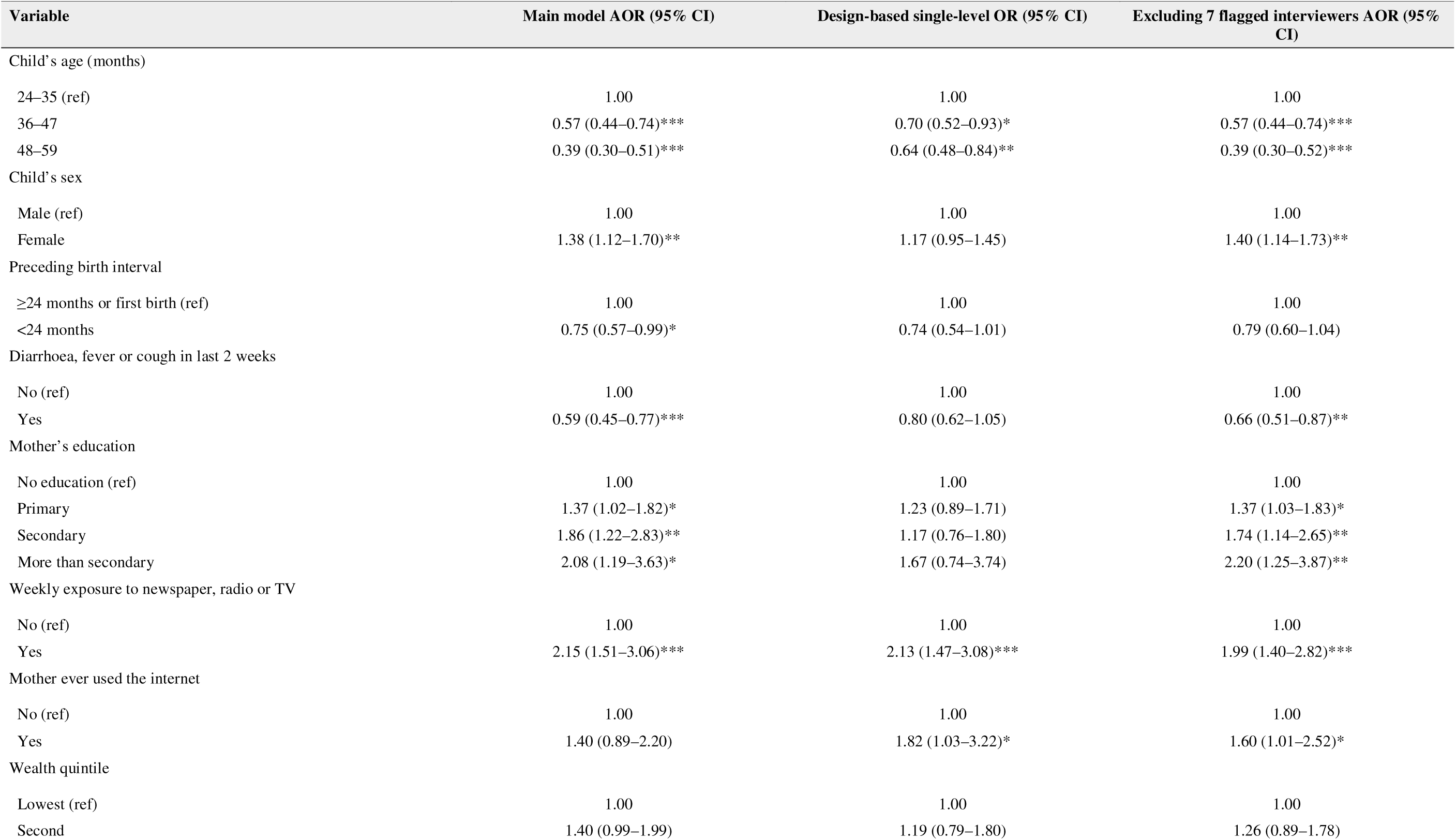
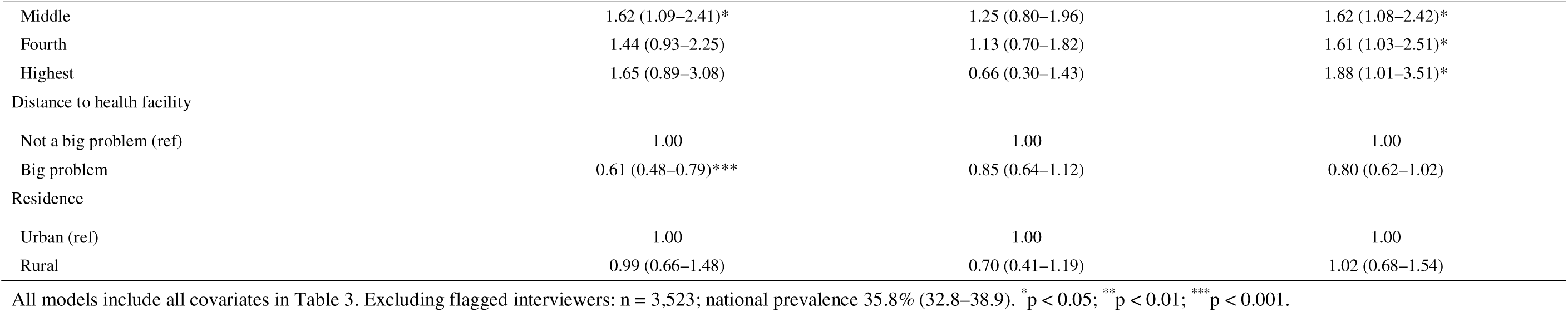
Sensitivity analyses: conditional (three-level) and population-averaged (design-based) odds ratios, and three-level model excluding flagged interviewers (selected variables)

| **Variable** | **Main model AOR (95% CI)** | **Design-based single-level OR (95% CI)** | **Excluding 7 flagged interviewers AOR (95% CI)** |
|---|---|---|---|
| Child’s age (months) |  |  |  |
| 24–35 (ref) | 1.00 | 1.00 | 1.00 |
| 36–47 | 0.57 (0.44–0.74)*** | 0.70 (0.52–0.93)* | 0.57 (0.44–0.74)*** |
| 48–59 | 0.39 (0.30–0.51)*** | 0.64 (0.48–0.84)** | 0.39 (0.30–0.52)*** |
| Child’s sex |  |  |  |
| Male (ref) | 1.00 | 1.00 | 1.00 |
| Female | 1.38 (1.12–1.70)** | 1.17 (0.95–1.45) | 1.40 (1.14–1.73)** |
| Preceding birth interval |  |  |  |
| ≥24 months or first birth (ref) | 1.00 | 1.00 | 1.00 |
| <24 months | 0.75 (0.57–0.99)* | 0.74 (0.54–1.01) | 0.79 (0.60–1.04) |
| Diarrhoea, fever or cough in last 2 weeks |  |  |  |
| No (ref) | 1.00 | 1.00 | 1.00 |
| Yes | 0.59 (0.45–0.77)*** | 0.80 (0.62–1.05) | 0.66 (0.51–0.87)** |
| Mother’s education |  |  |  |
| No education (ref) | 1.00 | 1.00 | 1.00 |
| Primary | 1.37 (1.02–1.82)* | 1.23 (0.89–1.71) | 1.37 (1.03–1.83)* |
| Secondary | 1.86 (1.22–2.83)** | 1.17 (0.76–1.80) | 1.74 (1.14–2.65)** |
| More than secondary | 2.08 (1.19–3.63)* | 1.67 (0.74–3.74) | 2.20 (1.25–3.87)** |
| Weekly exposure to newspaper, radio or TV |  |  |  |
| No (ref) | 1.00 | 1.00 | 1.00 |
| Yes | 2.15 (1.51–3.06)*** | 2.13 (1.47–3.08)*** | 1.99 (1.40–2.82)*** |
| Mother ever used the internet |  |  |  |
| No (ref) | 1.00 | 1.00 | 1.00 |
| Yes | 1.40 (0.89–2.20) | 1.82 (1.03–3.22)* | 1.60 (1.01–2.52)* |
| Wealth quintile |  |  |  |
| Lowest (ref) | 1.00 | 1.00 | 1.00 |
| Second | 1.40 (0.99–1.99) | 1.19 (0.79–1.80) | 1.26 (0.89–1.78) |
| Middle | 1.62 (1.09–2.41)* | 1.25 (0.80–1.96) | 1.62 (1.08–2.42)* |
| Fourth | 1.44 (0.93–2.25) | 1.13 (0.70–1.82) | 1.61 (1.03–2.51)* |
| Highest | 1.65 (0.89–3.08) | 0.66 (0.30–1.43) | 1.88 (1.01–3.51)* |
| Distance to health facility |  |  |  |
| Not a big problem (ref) | 1.00 | 1.00 | 1.00 |
| Big problem | 0.61 (0.48–0.79)*** | 0.85 (0.64–1.12) | 0.80 (0.62–1.02) |
| Residence |  |  |  |
| Urban (ref) | 1.00 | 1.00 | 1.00 |
| Rural | 0.99 (0.66–1.48) | 0.70 (0.41–1.19) | 1.02 (0.68–1.54) |

All models include all covariates in Table 3. Excluding flagged interviewers: n = 3,523; national prevalence 35.8% (32.8–38.9). <sup>\*</sup>p < 0.05; <sup>\*\*</sup>p < 0.01; <sup>\*\*\*</sup>p < 0.001.

**Additional file 1, Table S3.** Interviewers flagged for outlying proportions of children on track (clusterstratified Mantel–Haenszel test, two-sided Bonferroni p < 0.05/161)

| **Interviewer ID (v028)** | **Region** | **Children (n)** | **% on track** | **Expected<sup>a</sup>** | **Observed** | **z** |
|---|---|---|---|---|---|---|
| 3007 | Harari | 20 | 5.0 | 9.1 | 1 | -4.5 |
| 3706 | Dire Dawa | 25 | 16.0 | 10.9 | 4 | -3.9 |
| 2207 | Central Ethiopia | 41 | 7.3 | 11.0 | 3 | -3.8 |
| 2306 | Central Ethiopia | 38 | 89.5 | 25.6 | 34 | 3.8 |
| 1205 | South Ethiopia | 29 | 65.5 | 9.8 | 17 | 3.9 |
| 3707 | Dire Dawa | 31 | 87.1 | 10.1 | 18 | 4.4 |
| 3307 | South West Ethiopia | 24 | 83.3 | 9.1 | 18 | 5.0 |

<sup>a</sup> Expected number on track based on other interviewers working in the same clusters.

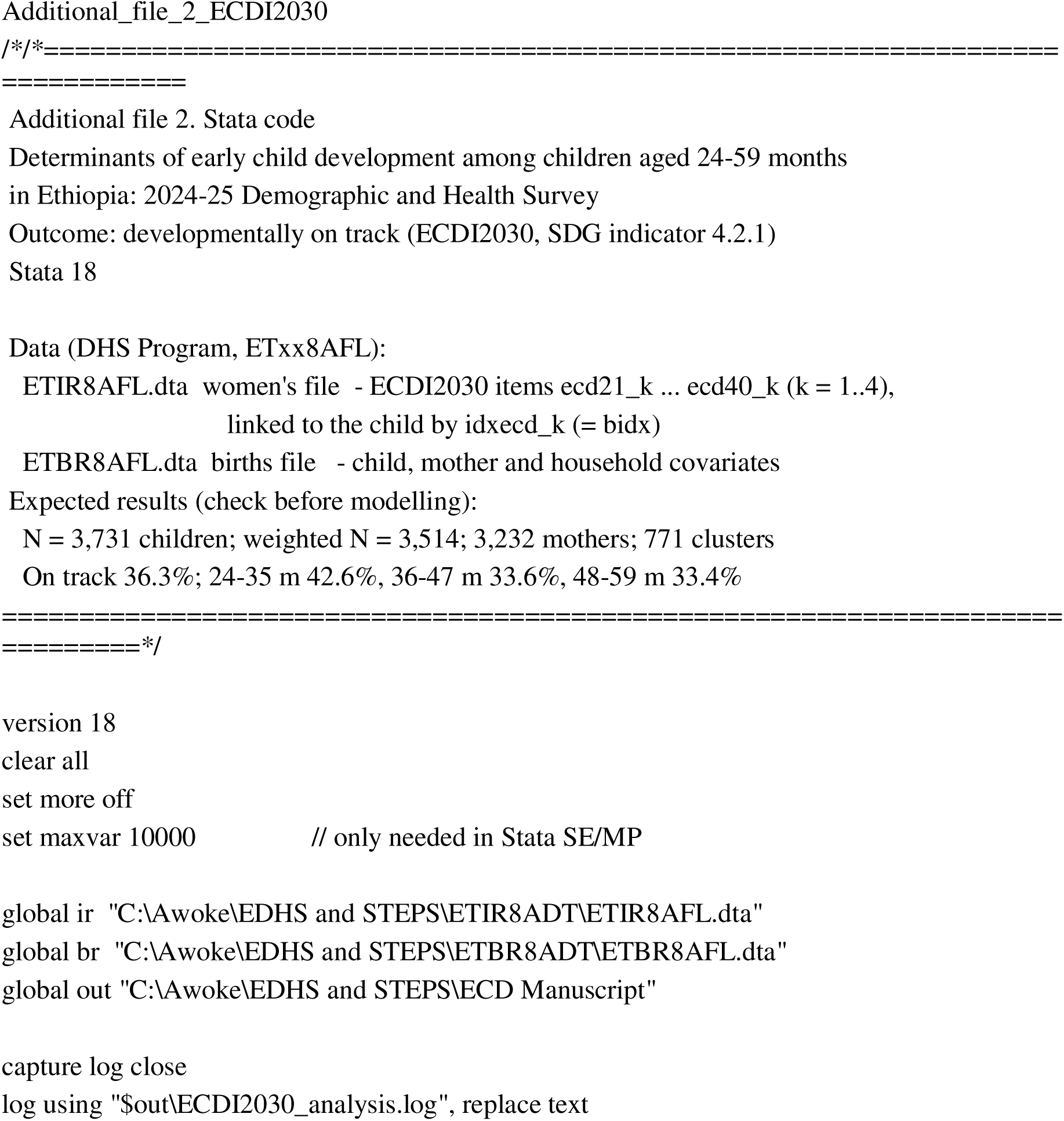
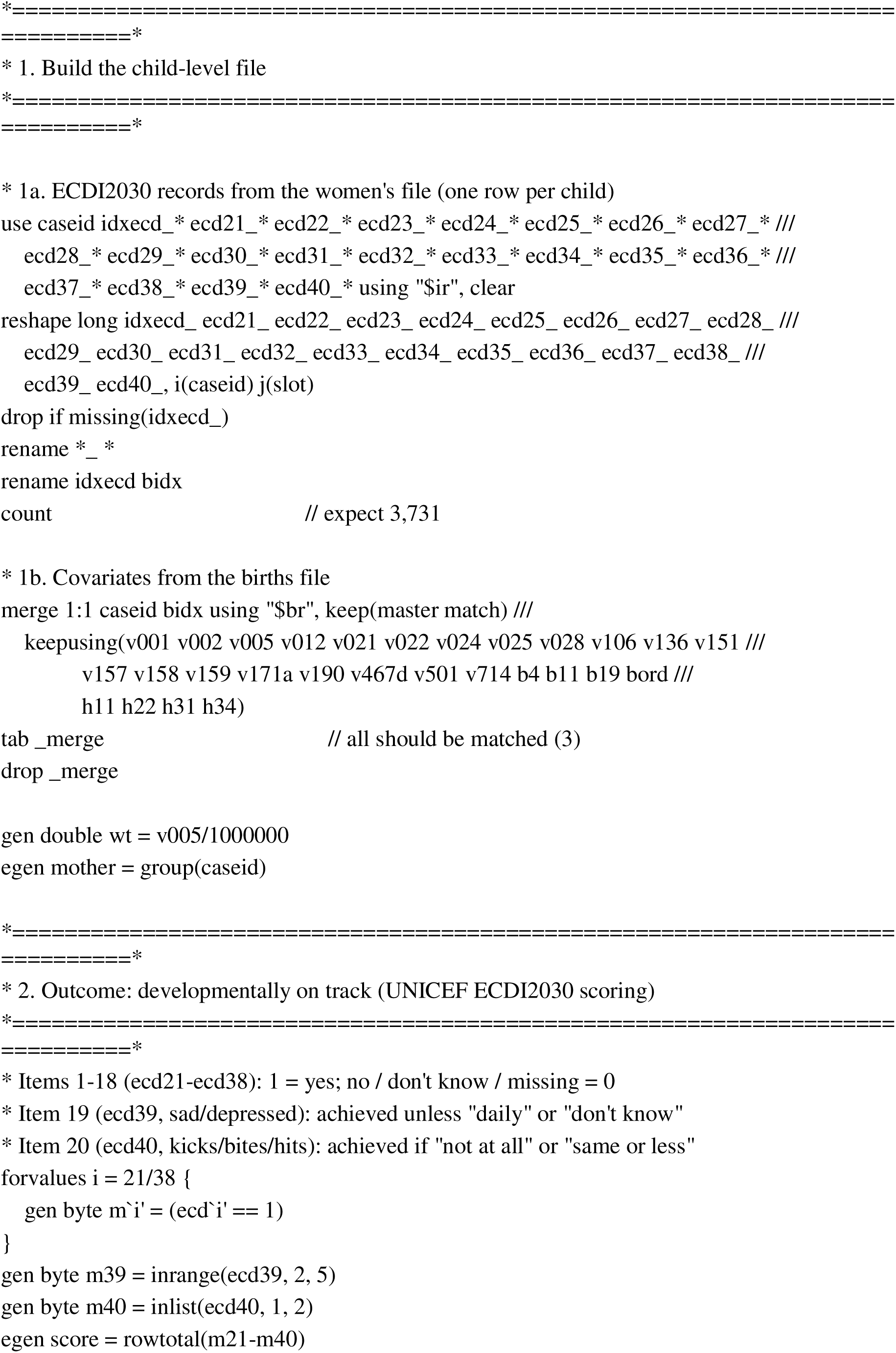
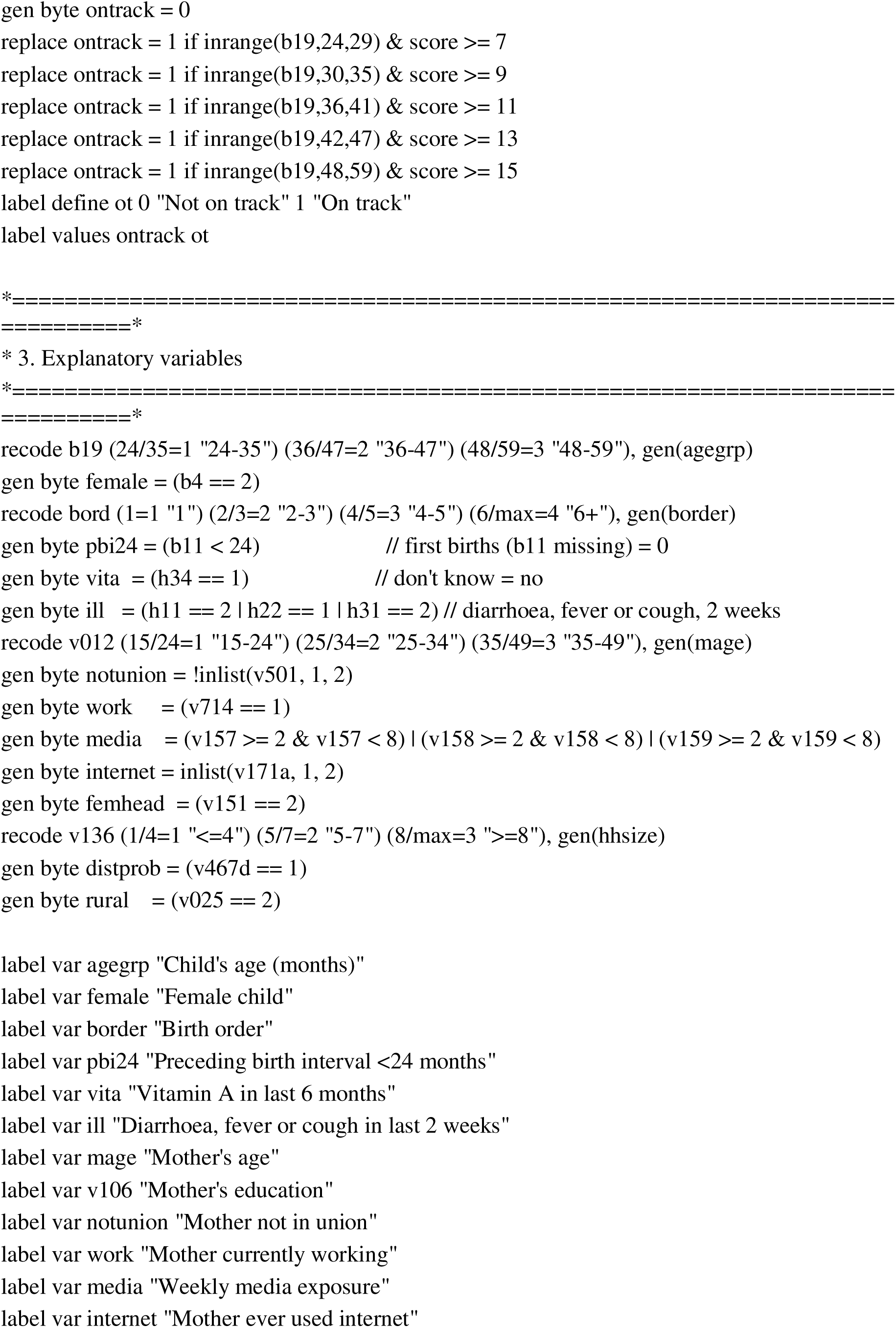
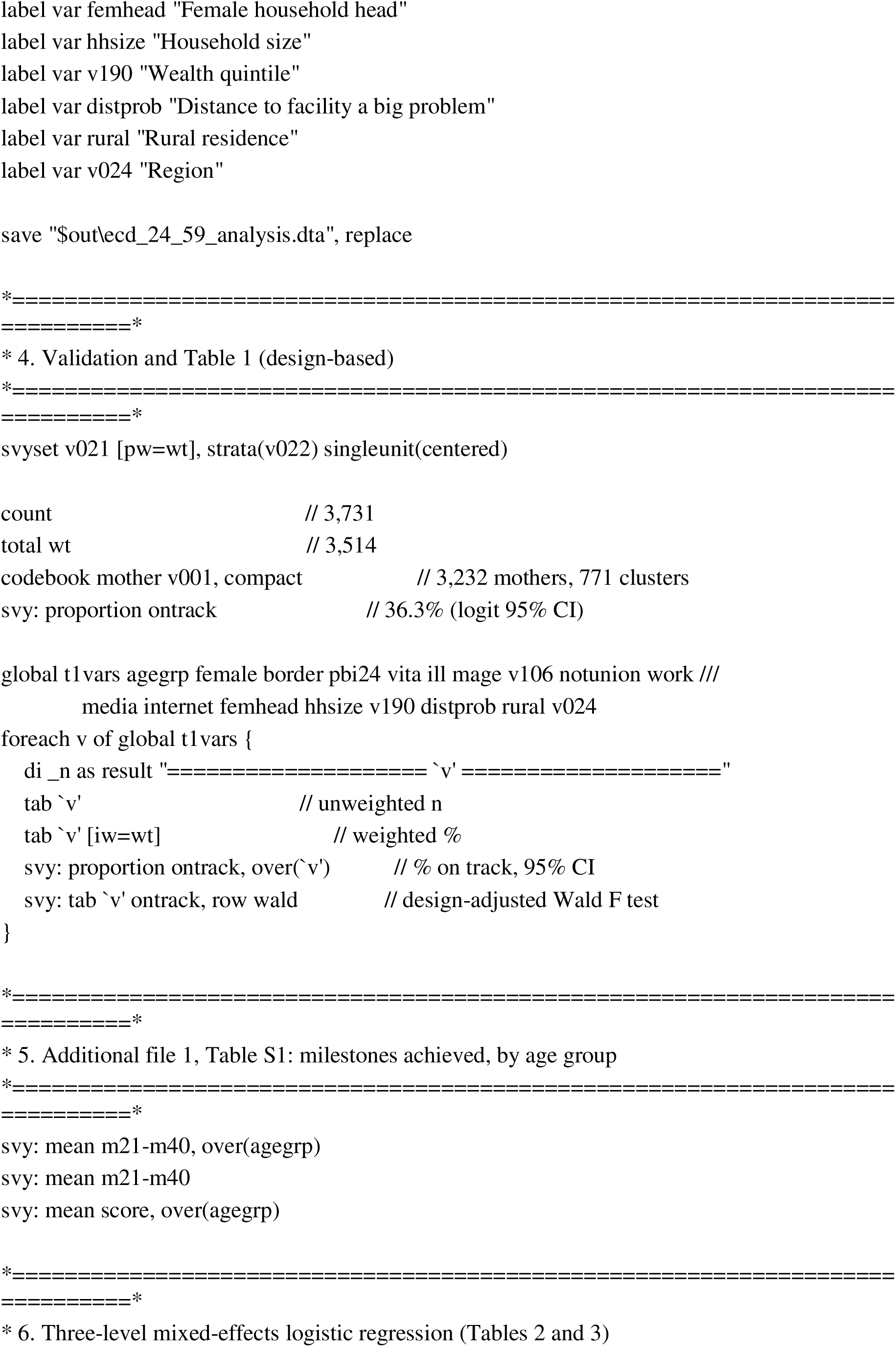
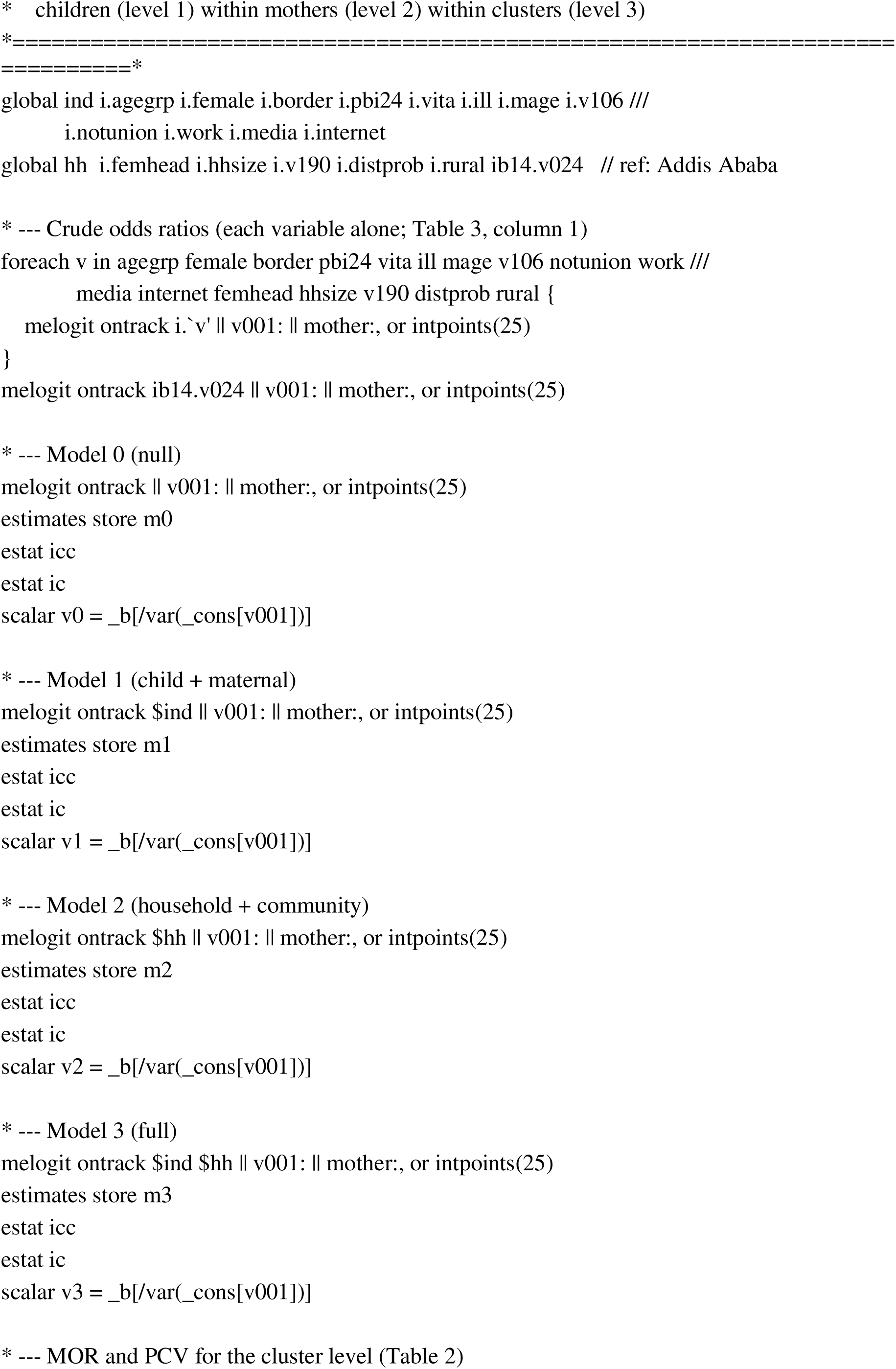
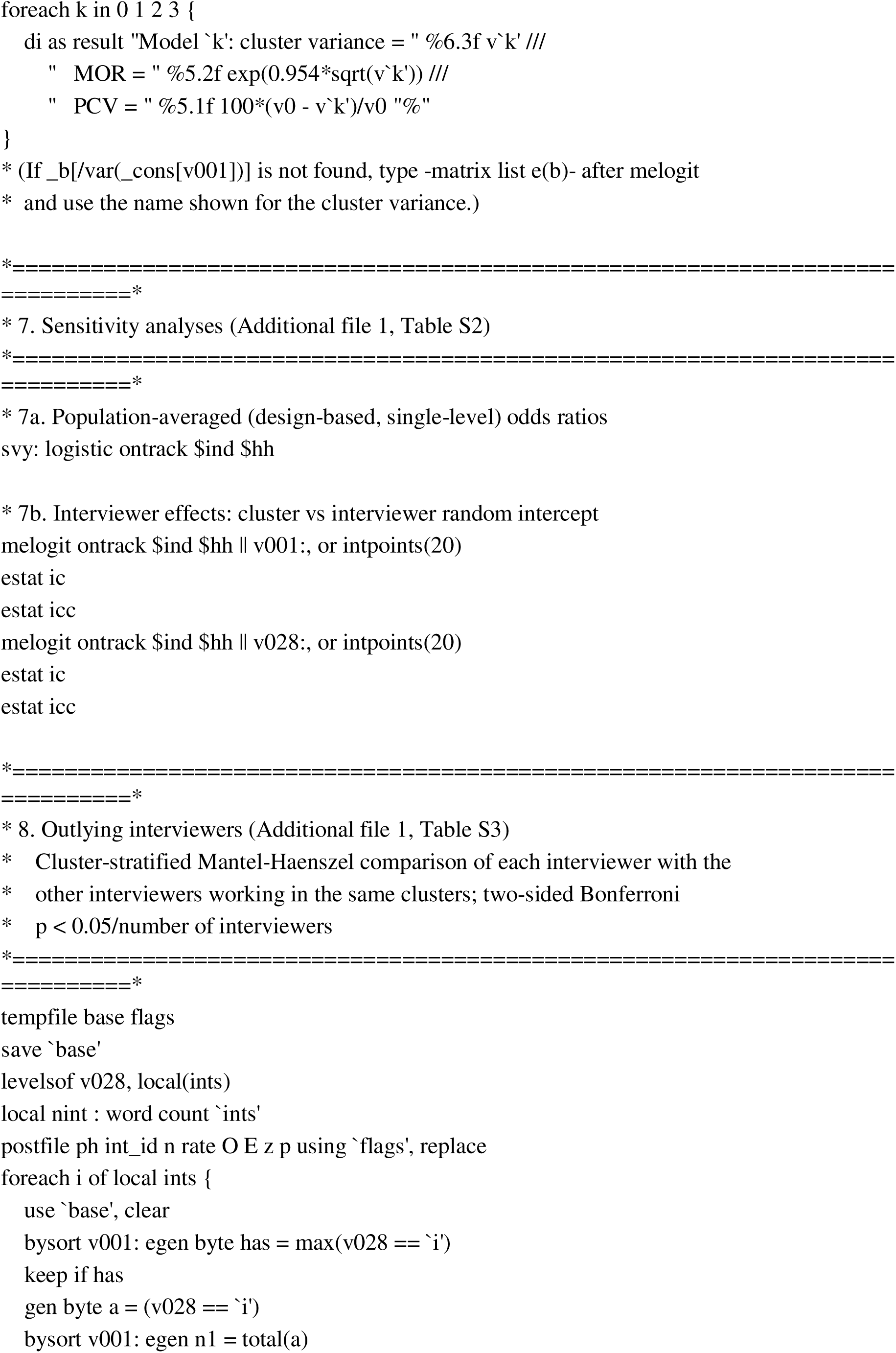
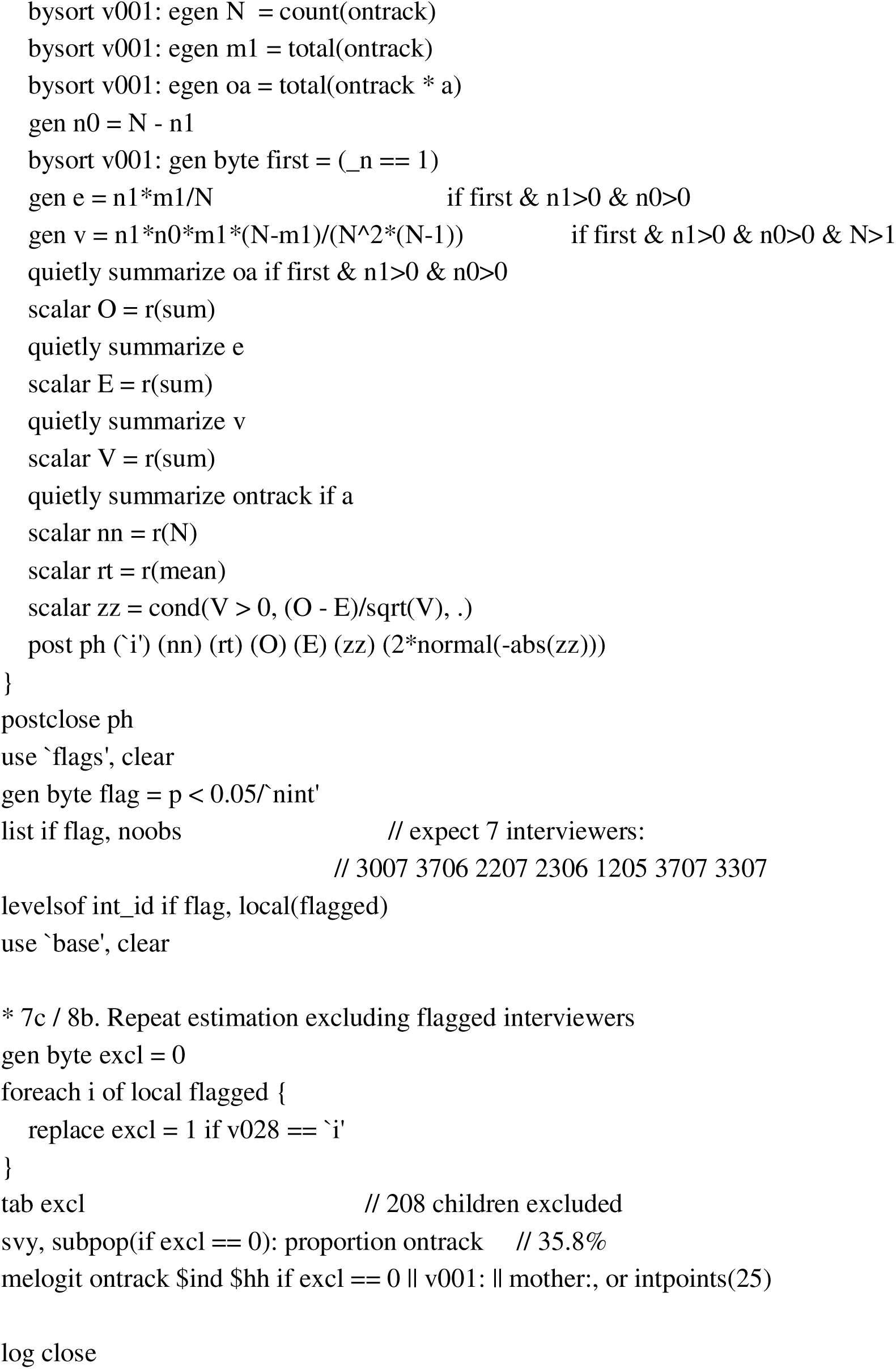

| Additional_file_2_ECDI2030 |
|---|
| /*/*============================================================================== |
| Additional file 2. Stata code |
| Determinants of early child development among children aged 24-59 months |
| in Ethiopia: 2024-25 Demographic and Health Survey |
| Outcome: developmentally on track (ECDI2030, SDG indicator 4.2.1) |
| Stata 18 |
| Data (DHS Program, ETxx8AFL): |
| ETIR8AFL.dta women's file - ECDI2030 items ecd21_k ... ecd40_k (k = 1..4), |
| linked to the child by idxecd_k (= bidx) |
| ETBR8AFL.dta births file - child, mother and household covariates |
| Expected results (check before modelling): |
| N = 3,731 children; weighted N = 3,514; 3,232 mothers; 771 clusters |
| On track 36.3%; 24-35 m 42.6%, 36-47 m 33.6%, 48-59 m 33.4% |
| =============================================================================*/ |
| version 18 |
| clear all |
| set more off |
| set maxvar 10000 // only needed in Stata SE/MP |
| global ir "C:\Awoke\EDHS and STEPS\ETIR8ADT\ETIR8AFL.dta" |
| global br "C:\Awoke\EDHS and STEPS\ETBR8ADT\ETBR8AFL.dta" |
| global out "C:\Awoke\EDHS and STEPS\ECD Manuscript" |
| capture log close |
| log using "\$out\ECDI2030_analysis.log", replace text |
| *=============================================================================* |
| * 1. Build the child-level file |
| *=============================================================================* |
| * 1a. ECDI2030 records from the women's file (one row per child) |
| use caseid idxecd_* ecd21_* ecd22_* ecd23_* ecd24_* ecd25_* ecd26_* ecd27_* /// |
| ecd28_* ecd29_* ecd30_* ecd31_* ecd32_* ecd33_* ecd34_* ecd35_* ecd36_* /// |
| ecd37_* ecd38_* ecd39_* ecd40_* using "\$ir", clear |
| reshape long idxecd_ ecd21_ ecd22_ ecd23_ ecd24_ ecd25_ ecd26_ ecd27_ ecd28_ /// |
| ecd29_ ecd30_ ecd31_ ecd32_ ecd33_ ecd34_ ecd35_ ecd36_ ecd37_ ecd38_ /// |
| ecd39_ ecd40_, i(caseid) j(slot) |
| drop if missing(idxecd_) |
| rename *_ * |
| rename idxecd bidx |
| count // expect 3,731 |
| * 1b. Covariates from the births file |
| merge 1:1 caseid bidx using "\$br", keep(master match) /// |
| keepusing(v001 v002 v005 v012 v021 v022 v024 v025 v028 v106 v136 v151 /// |
| v157 v158 v159 v171a v190 v467d v501 v714 b4 b11 b19 bord /// |
| h11 h22 h31 h34) |
| tab _merge // all should be matched (3) |
| drop _merge |
| gen double wt = v005/1000000 |
| egen mother = group(caseid) |
| *=============================================================================* |
| * 2. Outcome: developmentally on track (UNICEF ECDI2030 scoring) |
| *=============================================================================* |
| * Items 1-18 (ecd21-ecd38): 1 = yes; no / don't know / missing = 0 |
| * Item 19 (ecd39, sad/depressed): achieved unless "daily" or "don't know" |
| * Item 20 (ecd40, kicks/bites/hits): achieved if "not at all" or "same or less" |
| forvalues i = 21/38 { |
| gen byte m`i' = (ecd`i' == 1) |
| } |
| gen byte m39 = inrange(ecd39, 2, 5) |
| gen byte m40 = inlist(ecd40, 1, 2) |
| egen score = rowtotal(m21-m40) |
| gen byte ontrack = 0 |
| replace ontrack = 1 if inrange(b19,24,29) & score >= 7 |
| replace ontrack = 1 if inrange(b19,30,35) & score >= 9 |
| replace ontrack = 1 if inrange(b19,36,41) & score >= 11 |
| replace ontrack = 1 if inrange(b19,42,47) & score >= 13 |
| replace ontrack = 1 if inrange(b19,48,59) & score >= 15 |
| label define ot 0 "Not on track" 1 "On track" |
| label values ontrack ot |
| *=============================================================================* |
| * 3. Explanatory variables |
| *=============================================================================* |
| recode b19 (24/35=1 "24-35") (36/47=2 "36-47") (48/59=3 "48-59"), gen(agegrp) |
| gen byte female = (b4 == 2) |
| recode bord (1=1 "1") (2/3=2 "2-3") (4/5=3 "4-5") (6/max=4 "6+"), gen(border) |
| gen byte pbi24 = (b11 < 24) // first births (b11 missing) = 0 |
| gen byte vita = (h34 == 1) // don't know = no |
| gen byte ill = (h11 == 2 \| h22 == 1 \| h31 == 2) // diarrhoea, fever or cough, 2 weeks |
| recode v012 (15/24=1 "15-24") (25/34=2 "25-34") (35/49=3 "35-49"), gen(mage) |
| gen byte notunion = !inlist(v501, 1, 2) |
| gen byte work = (v714 == 1) |
| gen byte media = (v157 >= 2 & v157 < 8) \| (v158 >= 2 & v158 < 8) \| (v159 >= 2 & v159 < 8) |
| gen byte internet = inlist(v171a, 1, 2) |
| gen byte femhead = (v151 == 2) |
| recode v136 (1/4=1 "<=4") (5/7=2 "5-7") (8/max=3 ">=8"), gen(hhsize) |
| gen byte distprob = (v467d == 1) |
| gen byte rural = (v025 == 2) |
| label var agegrp "Child's age (months)" |
| label var female "Female child" |
| label var border "Birth order" |
| label var pbi24 "Preceding birth interval <24 months" |
| label var vita "Vitamin A in last 6 months" |
| label var ill "Diarrhoea, fever or cough in last 2 weeks" |
| label var mage "Mother's age" |
| label var v106 "Mother's education" |
| label var notunion "Mother not in union" |
| label var work "Mother currently working" |
| label var media "Weekly media exposure" |
| label var internet "Mother ever used internet" |
| label var femhead "Female household head" |
| label var hhsize "Household size" |
| label var v190 "Wealth quintile" |
| label var distprob "Distance to facility a big problem" |
| label var rural "Rural residence" |
| label var v024 "Region" |
| save "\$out\ecd_24_59_analysis.dta", replace |
| *=============================================================================* |
| * 4. Validation and Table 1 (design-based) |
| *=============================================================================* |
| svyset v021 [pw=wt], strata(v022) singleunit(centered) |
| count // 3,731 |
| total wt // 3,514 |
| codebook mother v001, compact // 3,232 mothers, 771 clusters |
| svy: proportion ontrack // 36.3% (logit 95% CI) |
| global t1vars agegrp female border pbi24 vita ill mage v106 notunion work /// |
| media internet femhead hhsize v190 distprob rural v024 |
| foreach v of global t1vars { |
| di _n as result "==================== `v' ====================" |
| tab `v' // unweighted n |
| tab `v' [iw=wt] // weighted % |
| svy: proportion ontrack, over(`v') // % on track, 95% CI |
| svy: tab `v' ontrack, row wald // design-adjusted Wald F test |
| } |
| *=============================================================================* |
| * 5. Additional file 1, Table S1: milestones achieved, by age group |
| *=============================================================================* |
| svy: mean m21-m40, over(agegrp) |
| svy: mean m21-m40 |
| svy: mean score, over(agegrp) |
| *=============================================================================* |
| * 6. Three-level mixed-effects logistic regression (Tables 2 and 3) |
| * children (level 1) within mothers (level 2) within clusters (level 3) |
| *=============================================================================* |
| global ind i.agegrp i.female i.border i.pbi24 i.vita i.ill i.mage i.v106 /// |
| i.notunion i.work i.media i.internet |
| global hh i.femhead i.hhsize i.v190 i.distprob i.rural ib14.v024 // ref: Addis Ababa |
| * --- Crude odds ratios (each variable alone; Table 3, column 1) |
| foreach v in agegrp female border pbi24 vita ill mage v106 notunion work /// |
| media internet femhead hhsize v190 distprob rural { |
| melogit ontrack i.`v' \|\| v001: \|\| mother:, or intpoints(25) |
| } |
| melogit ontrack ib14.v024 \|\| v001: \|\| mother:, or intpoints(25) |
| * --- Model 0 (null) |
| melogit ontrack \|\| v001: \|\| mother:, or intpoints(25) |
| estimates store m0 |
| estat icc |
| estat ic |
| scalar v0 = _b[/var(_cons[v001])] |
| * --- Model 1 (child + maternal) |
| melogit ontrack \$ind \|\| v001: \|\| mother:, or intpoints(25) |
| estimates store m1 |
| estat icc |
| estat ic |
| scalar v1 = _b[/var(_cons[v001])] |
| * --- Model 2 (household + community) |
| melogit ontrack \$hh \|\| v001: \|\| mother:, or intpoints(25) |
| estimates store m2 |
| estat icc |
| estat ic |
| scalar v2 = _b[/var(_cons[v001])] |
| * --- Model 3 (full) |
| melogit ontrack \$ind \$hh \|\| v001: \|\| mother:, or intpoints(25) |
| estimates store m3 |
| estat icc |
| estat ic |
| scalar v3 = _b[/var(_cons[v001])] |
| * --- MOR and PCV for the cluster level (Table 2) |
| foreach k in 0 1 2 3 { |
| di as result "Model `k': cluster variance = " %6.3f v`k' /// |
| " MOR = " %5.2f exp(0.954*sqrt(v`k')) /// |
| " PCV = " %5.1f 100*(v0 - v`k')/v0 "%" |
| } |
| * (If _b[/var(_cons[v001])] is not found, type -matrix list e(b)- after melogit |
| * and use the name shown for the cluster variance.) |
| *=============================================================================* |
| * 7. Sensitivity analyses (Additional file 1, Table S2) |
| *=============================================================================* |
| * 7a. Population-averaged (design-based, single-level) odds ratios |
| svy: logistic ontrack \$ind \$hh |
| * 7b. Interviewer effects: cluster vs interviewer random intercept |
| melogit ontrack \$ind \$hh \|\| v001:, or intpoints(20) |
| estat ic |
| estat icc |
| melogit ontrack \$ind \$hh \|\| v028:, or intpoints(20) |
| estat ic |
| estat icc |
| *=============================================================================* |
| * 8. Outlying interviewers (Additional file 1, Table S3) |
| * Cluster-stratified Mantel-Haenszel comparison of each interviewer with the |
| * other interviewers working in the same clusters; two-sided Bonferroni |
| * p < 0.05/number of interviewers |
| *=============================================================================* |
| tempfile base flags |
| save `base' |
| levelsof v028, local(ints) |
| local nint : word count `ints' |
| postfile ph int_id n rate O E z p using `flags', replace |
| foreach i of local ints { |
| use `base', clear |
| bysort v001: egen byte has = max(v028 == `i') |
| keep if has |
| gen byte a = (v028 == `i') |
| bysort v001: egen n1 = total(a) |
| bysort v001: egen N = count(ontrack) |
| bysort v001: egen m1 = total(ontrack) |
| bysort v001: egen oa = total(ontrack * a) |
| gen n0 = N - n1 |
| bysort v001: gen byte first = (_n == 1) |
| gen e = n1*m1/N if first & n1>0 & n0>0 |
| gen v = n1*n0*m1*(N-m1)/(N^2*(N-1)) if first & n1>0 & n0>0 & N>1 |
| quietly summarize oa if first & n1>0 & n0>0 |
| scalar O = r(sum) |
| quietly summarize e |
| scalar E = r(sum) |
| quietly summarize v |
| scalar V = r(sum) |
| quietly summarize ontrack if a |
| scalar nn = r(N) |
| scalar rt = r(mean) |
| scalar zz = cond(V > 0, (O - E)/sqrt(V), .) |
| post ph (`i') (nn) (rt) (O) (E) (zz) (2*normal(-abs(zz))) |
| } |
| postclose ph |
| use `flags', clear |
| gen byte flag = p < 0.05/`nint' |
| list if flag, noobs // expect 7 interviewers: |
| // 3007 3706 2207 2306 1205 3707 3307 |
| levelsof int_id if flag, local(flagged) |
| use `base', clear |
| * 7c / 8b. Repeat estimation excluding flagged interviewers |
| gen byte excl = 0 |
| foreach i of local flagged { |
| replace excl = 1 if v028 == `i' |
| } |
| tab excl // 208 children excluded |
| svy, subpop(if excl == 0): proportion ontrack // 35.8% |
| melogit ontrack \$ind \$hh if excl == 0 \|\| v001: \|\| mother:, or intpoints(25) |
| log close |

